# Length of Stay in Acute Burn Patients Treated with Intact Fish Skin Graft Versus Synthetic Skin Substitutes

**DOI:** 10.64898/2026.04.14.26350896

**Authors:** Rajiv Sood, Nathanael D. Hevelone, Ólafur B. Davíðsson, Ragnar P. Kristjánsson, Bart D. Phillips, John C. Lantis, Gunnar Johannsson

## Abstract

**Objective:** To compare hospital length of stay (LOS) and complication rates between patients treated with intact fish skin graft (IFSG; Kerecis) and synthetic/biosynthetic dermal substitutes (SSS; Integra and NovoSorb BTM) prior to autograft using the ABA Burn Care Quality Platform.

**Methods:** Adult acute-burn patients treated with either IFSG or SSS were identified from the American Burn Association Burn Care Quality Platform. IFSG patients were 1:4 propensity-matched to SSS patients on age, sex, TBSA, burn severity, inhalation injury, and trauma. Primary outcome was hospital length of stay (Gamma GLMM); composite secondary was sepsis, graft loss, VTE, or HAPI (binomial GLMM).

**Results:** 93 IFSG and 372 SSS patients across 49 burn centers were analyzed with acute burns represented as moderate-sized (mean TBSA 12.6%), with burn depth predominantly mixed or third-degree. GLMM-adjusted mean length of stay was 22.7 days (95% CI 18.2–28.3) for IFSG versus 35.2 days (95% CI 30.9–40.1) for SSS (ratio 0.646; p = 0.000235), a 12.4-day reduction. The composite complication rate was 7.5% (7/93) in IFSG versus 15.9% (59/372) in SSS (GLMM odds ratio 2.37, 95% CI 1.03–5.42; p = 0.041). All individual complications were numerically lower in the IFSG arm.

**Conclusions:** In this propensity-matched analysis of the ABA registry, IFSG was associated with a statistically significant 12.4-day reduction in length of stay and a significantly lower composite complication rate. IFSG is an effective alternative for burns requiring dermal substitution and autografting. Prospective studies in larger burns are warranted.

## Introduction

Acute burn injuries requiring surgical management remain a significant source of morbidity, prolonged hospitalization, and healthcare expenditure in the United States.^1^ Split-thickness skin grafting (STSG) continues to be the definitive treatment for deep partial-thickness and full-thickness burns; however, many patients require temporary wound coverage prior to autografting to prepare the wound bed, manage infection risk, and optimize conditions for graft take.^2^ Dermal substitutes and temporary wound coverage products serve this critical bridging function, yet the choice among available options remains guided more by institutional tradition or preference than by comparative effectiveness data from large patient populations.^3^

Several dermal regeneration scaffolds are available for wound bed preparation in deep burns. These products share a common functional goal: to provide a three-dimensional matrix for infiltration by host fibroblasts and endothelial cells, allowing for the formation of an organized neodermis capable of supporting subsequent STSG or, in some cases, direct epithelialization.^4,5^ The products differ primarily in scaffold composition and the timeline required for vascular integration.^4,5^

Synthetic and biosynthetic scaffolds including Integra Dermal Regeneration Template (IDRT) (Integra LifeSciences, Princeton, NJ) and NovoSorb Biodegradable Temporizing Matrix (BTM) (PolyNovo, Port Melbourne, Australia) employ bilayer constructs with a dermal regeneration layer (collagen/glycosaminoglycan matrix, approximately 0.8–1.0 mm for Integra; polyurethane foam, approximately 2.0 mm for NovoSorb) and a temporary outer barrier (silicone or sealing membrane) that is removed once vascular integration of the scaffold is complete. Published vascular integration times range from approximately 14 to 21 days for Integra^5^ and 15 to 128 days (with a median of 36 days) for NovoSorb.^4^ Reported subsequent autograft loss rates include approximately 12.3% for Integra^7^ and 15.5% for NovoSorb.^6^ Intact fish skin graft (IFSG; Graftguide, Kerecis, Arlington, VA) is a biologically derived dermal regeneration graft from North Atlantic cod (Gadus Morhua) that retains the native collagen architecture and omega-3 polyunsaturated fatty acid content of the source tissue.^8^ At approximately 0.7 mm thickness, the intact fish skin scaffold is fully incorporated into the wound bed without a removable barrier layer, functioning as a single-stage dermal regeneration graft. Preclinical and clinical evidence indicates that IFSG accelerates neovascularization and supports organized tissue regeneration with reduced contraction.^9–11^ Histologic analysis has demonstrated formation of a well-organized, vascularized neodermis, termed “keratinocyte-ready dermis” (KereDerm), that supports either STSG take or direct keratinocyte migration.^12^ IFSG has seen clinical utilization across a spectrum of burn depths, including deep partial-thickness wounds where the goal may be to promote epithelialization and reduce the need for autografting, and full-thickness burns requiring dermal regeneration prior to STSG.^11,13–16^ The former clinical use is the subject of an ongoing prospective randomized controlled trial (NCT07326657) and the focus of the present study. Despite substantial and growing clinical utilization of IFSG as a bridge-to-autograft, no large-scale registry comparison against established synthetic/biosynthetic dermal scaffolds has been performed.

Hospital length of stay is a critical outcome in burn care that extends well beyond resource utilization. Each additional day of hospitalization increases cumulative nosocomial infection, venous thromboembolism (VTE), hospital-acquired pressure injury, deconditioning, and psychological distress.^17^ Daily costs in specialized burn units in the united states average approximately $9,000,^18^ meaning that even modest LOS reductions carry substantial economic implications for healthcare systems and payers. If one category of dermal substitute enables earlier definitive closure without increasing complication rates, the clinical and economic consequences are significant and extend far beyond a simple reflection of product integration mechanics.

The objective of this study was to compare hospital length of stay and complication rates between IFSG and synthetic/biosynthetic dermal substitutes in propensity score–matched adult burn patients identified from the American Burn Association (ABA) Burn Care Quality Platform, a national registry representing the largest collection of burn center outcomes data in the United States.

## Methods

### Data Source and Independence of Analysis

This study used de-identified patient data from the American Burn Association Burn Care Quality Platform (BCQP), a registry that collects standardized clinical and outcome data from a network of approximately 110 burn centers across the United States and Canada. The study was funded by Kerecis LLC. Analyses were performed according to a pre-specified statistical analysis plan (SAP), which documented endpoints, matching covariates, and model types used for this report. The SAP was developed jointly by BData and Frameshift biostatisticians, with Frameshift contributing the analytic specification and method selection; BData independently performed all data extraction, propensity-score matching, and statistical analysis. The sponsor did not have access to patient-level data at any point during the study. Ethical review was performed by WCG IRB (ref. 20261461), which determined the study exempt under 45 CFR 46.104(d)(4) for secondary analysis of de-identified data.

### Study Population

Adult patients (aged 18 years or older) admitted to a burn center for treatment of acute thermal burn injury between 2019 and 2025 were eligible for inclusion (Figure 1). The BCQP was queried for cases involving intact fish skin graft (IFSG; Kerecis GraftGuide) or synthetic/biosynthetic dermal substitutes (NovoSorb Biodegradable Temporizing Matrix, BTM, or Integra Dermal Regeneration Template, IDRT), with a documented subsequent autograft procedure during the same hospital admission. To minimize the influence of implausible hospitalizations likely to be data-entry errors, cases with recorded length of stay greater than 120 days were excluded. To reduce confounding, the analytic cohort was further restricted to patients who received either IFSG or SSS (not both) and who did not receive allograft (a pure-case cohort). After these exclusions, 102 IFSG-treated cases and 730 SSS-treated cases were available for matching (Figure 1).

**Figure 1.**
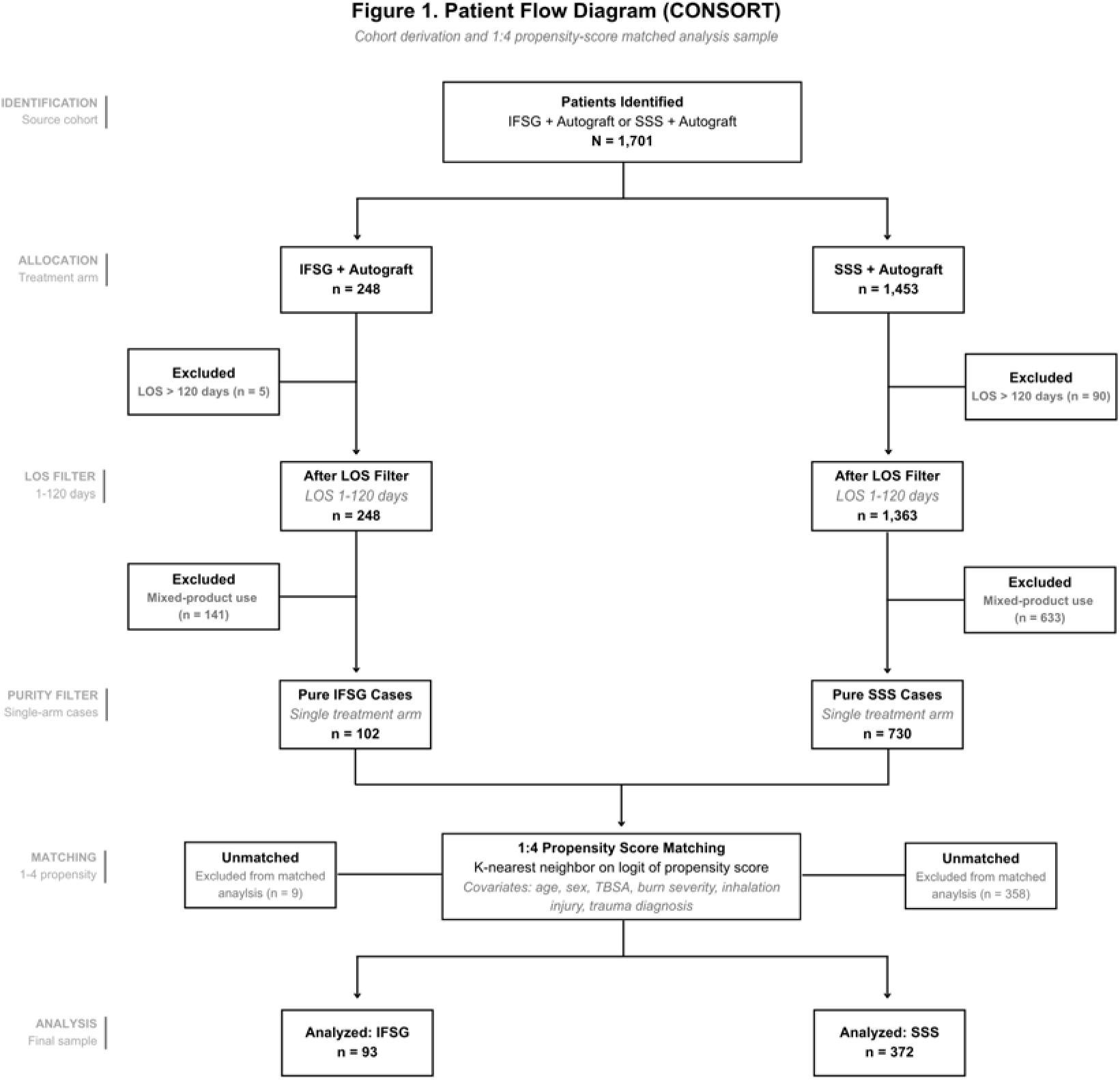
Patient flow diagram. From the ABA Burn Care Quality Platform, 248 patients received IFSG and autograft and 1,453 patients received SSS and autograft. After excluding 5 IFSG patients with a LOS >120 and 90 SSS patients with a LOS >120, 243 IFSG patients and 1363 SSS patients remained. Next, a subset was taken of the cases with a single treatment arm only (i.e. IFSG + autograft without SSS or allograft OR SSS + autograft without IFSG or allograft), 102 IFSG patients and 730 SSS patients remained. 93 IFSG patients were then matched 1:4 to 372 SSS patients from a pool of 730 SSS-eligible patients.

### Defining the Final IFSG and SSS Cohorts Through Propensity Matching

The American Burn Association Burn Care Quality Platform (BCQP) assigns products to product-category level. Within the registry, “Synthetic and Biosynthetic Skin Substitutes” (SSS) is captured as a pooled category that encompasses commercially available acellular and cellular biosynthetic constructs used for dermal substitution in acute burn care (e.g., bilayer collagen-glycosaminoglycan matrices, polyurethane-based biodegradable). Because the registry does not permit identification of specific synthetic or biosynthetic products, all comparisons in this study were performed at the category level (IFSG versus pooled SSS). IFSG and SSS cases were matched on propensity score estimated by logistic regression. Matching covariates were patient age, sex, percent total body surface area burned (TBSA), presence or absence of concomitant trauma diagnosis, presence or absence of concomitant inhalation injury, and burn depth classification (second-degree only, third-degree only, or mixed second and third degree). Matching was performed in Python using the PsmPy library, with K-nearest-neighbor matching on the logit of the propensity score, without replacement and without a caliper, at a 1:4 ratio (IFSG to SSS). The 1:4 ratio was selected during pre-match exploration to balance covariate balance against retained sample size and was fixed prior to outcome analysis. The random seed was fixed for reproducibility. After matching, the analytic cohort comprised 93 IFSG cases and 372 SSS cases (Figure 1).

Post-match covariate balance was assessed using absolute standardized mean differences (SMDs). An SMD of less than 0.10 was considered to indicate good balance, consistent with the conventional threshold.^19,20^

### Primary Endpoint

The primary outcome was hospital length of stay (LOS), defined as the number of days from admission to discharge.

### Secondary Endpoints

The pre-specified secondary endpoints were four BCQP-standardized in-hospital complications, each recognized in the burn literature as a major contributor to prolonged hospitalization, morbidity, and mortality: severe sepsis or septic shock, graft loss requiring repeat procedure, venous thromboembolism (deep vein thrombosis or pulmonary embolism), and hospital-acquired pressure injury (HAPI). Component-level definitions followed the BCQP standard definitions, and each endpoint was analyzed individually. In addition, we report a post-hoc composite of these four pre-specified events, defined as a binary indicator of whether the patient experienced any of severe sepsis or septic shock, graft loss requiring repeat procedure, venous thromboembolism, or HAPI. The composite was specified to provide a single, clinically interpretable, patient-centered measure of serious in-hospital morbidity in a population in which any one of these events would be expected to materially prolong burn hospitalization.

### Statistical Analysis

LOS was modelled using a generalized linear mixed model (GLMM) with a Gamma distribution and log-link function. Treatment arm was modelled as a two-level factor with IFSG as the reference level. Random intercepts for facility and for matched set were included to account for between-center variation and within-matched-set correlation. Estimated marginal means with 95% confidence intervals were obtained from the fitted model, and the pairwise ratio (IFSG to SSS) is reported with a nominal p-value. With a single pairwise comparison in the two-arm design, multiplicity adjustment was not applicable.

The composite secondary endpoint was analyzed using a binomial GLMM with a logit link, treatment arm as a two-level factor (IFSG reference), and a random intercept for matched set. Odds ratios (SSS vs IFSG) with 95% confidence intervals are reported. Two prespecified sensitivity analyses were performed: a binomial-logit GLMM with an additional random intercept for facility, and conditional logistic regression stratified on matched set.

The four constituent complication endpoints (sepsis, graft loss, VTE, HAPI) were analyzed individually using the same binomial-logit GLMM specification with a matched-set random effect. Event counts were low (single digits to low double digits per arm), and the resulting odds-ratio estimates have wide confidence intervals; the individual-endpoint results should be interpreted as directional rather than precise. Unadjusted comparisons used Fisher’s exact test.

Analyses were performed in R (R Foundation for Statistical Computing) using the glmmTMB, emmeans, lme4, and survival packages, and in Python using the PsmPy library for propensity-score matching. Statistical significance was defined as p < 0.05 (two-sided).

### Sensitivity Analyses

A prespecified sensitivity analysis for the primary endpoint was performed in a broader mixed-product cohort that included patients who received IFSG alongside other dermal substitutes including SSS or allograft during the same hospitalization. In this expanded cohort, 229 IFSG patients (with or without SSS or allograft) were matched 1:2 to 458 SSS patients (with or without allograft but without IFSG) across 67 burn centers using the same GLMM methodology, to evaluate the robustness of the primary finding across cohort definitions.

## Results

### IFSG-Treated and SSS-Treated Cohorts Were Well-Balanced

Propensity score matching was successful in balancing the IFSG and SSS treatment groups in that standardized mean differences between the two groups were reduced to <0.1 (and most <0.05) for all covariates (Figure 2). Covariates were either exactly balanced or nearly balanced; for those nearly balanced, the differences between groups were not statistically significant (Table 1).

**Figure 2.**
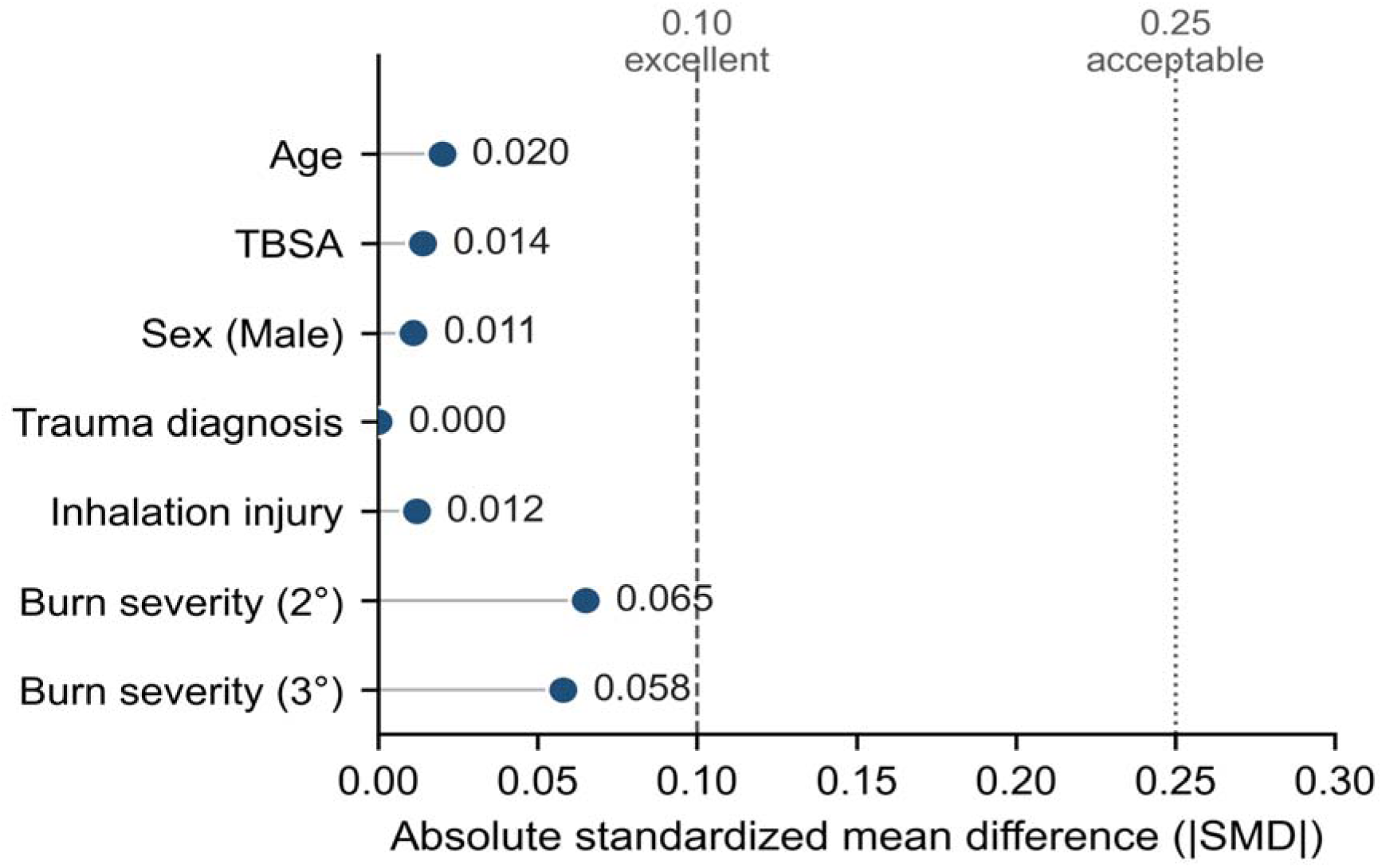
Post-match covariate balance assessed by absolute standardized mean differences (|SMD|). All seven matching covariates achieved balance below the 0.10 excellent threshold; the maximum absolute SMD was 0.065 (burn severity, second-degree). Dashed line denotes the 0.10 excellent balance threshold; dotted line denotes the 0.25 acceptable threshold.

**Table 1.** Baseline Demographic and Injury Characteristics of Propensity Score–Matched Cohorts.

| Characteristic | IFSG<br>(n = 93) | SSS<br>(n = 372) | SMD |
| --- | --- | --- | --- |
| Age, mean ± SD (years) | 50.68 ± 17.4 | 50.37 ± 17.4 | 0.017 |
| Male sex, n (%) | 56 (60.2) | 222 (59.7) | 0.011 |
| TBSA, mean ± SD (%) | 12.49 ± 12.6 | 12.67 ± 13.1 | 0.014 |
| Inhalation injury, n (%) | 5 (5.4) | 19 (5.1) | 0.012 |
| Trauma diagnosis, n (%) | 11 (11.8) | 44 (11.8) | 0.000 |
| **Burn severity classification** |  |  |  |
| 2nd degree only, n (%) | 29 (31.2) | 105 (28.2) | 0.065 |
| 3rd degree only, n (%) | 22 (23.7) | 79 (21.2) | 0.058 |
| Mixed depth, n (%) | 42 (45.2) | 188 (50.5) | 0.108 |

SMD = standardized mean difference, computed from post-match marginal means/standard deviations (continuous) or proportions (binary). All SMDs < 0.25 indicate acceptable balance; SMDs < 0.10 indicate excellent balance (Austin, 2009). IFSG = Kerecis Omega3 Burn. SSS = Integra DRT + NovoSorb BTM.

The mean patient ages were 50.7 years and 50.4 years for the IFSG-treated group and SSS-treated group, respectively (Table 1). Both groups were approximately 60% male. TBSA averaged 12.5% for the IFSG-treated group and 12.7% for the SSS-treated group. Concomitant trauma was present in 12% of patients in both groups and concomitant inhalation injury was present in 5% of patients in both groups. Approximately half of patients had both 2nd and 3rd degree burns. Patients with only 2nd degree burns made up 31% and 28% of the IFSG-treated group and SSS-treated group, respectively. Patients with only 3rd degree burns comprised 24% and 21% of the IFSG-treated group and SSS-treated group, respectively. Notably, the IFSG arm carried a higher diabetes burden than the SSS arm (30.1% vs 21.2%). Diabetes was not a matching covariate; this imbalance is addressed in the Discussion.

### Hospital Length-of-Stay Was Shorter in the IFSG-Treated Cohort

Unadjusted mean LOS was 24.1 days (SD 17.4; median 20, IQR 11–32) in the IFSG-treated group and 36.7 days (SD 25.1; median 31, IQR 17–52) in the SSS-treated group (Table 2, Figure 3), representing an unadjusted difference of 12.6 days that was statistically significant (p= 5.48e-08). To control for random effects causing clustering at the burn center-level and correlation within matched pairs, LOS was calculated after adjusting for these factors with a GLMM. Adjusted estimated marginal mean LOS was 22.7 days (95% CI, 18.2 – 28.3) in the IFSG-treated group and 35.2 days (95% CI, 30.9 – 40.1) in the SSS-treated group (Table 2, Figure 4). Thus, the adjusted LOS was 12.4 days longer on average in the SSS-treated group, which reached statistical significance (ratio 0.646; p = 0.000235).

**Figure 3.**
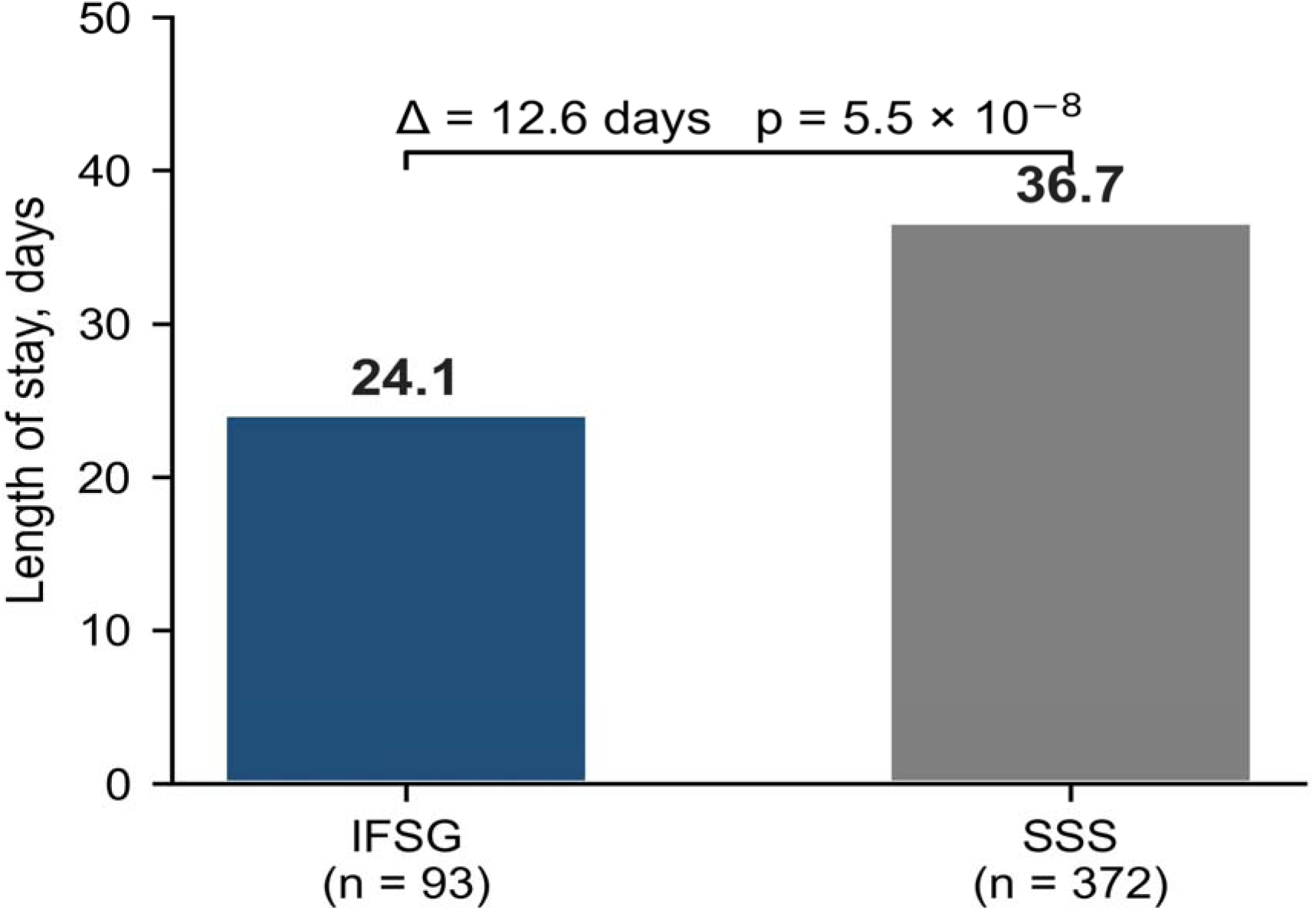
Unadjusted hospital length of stay. Mean LOS was 24.1 days in the IFSG-treated arm (n = 93) versus 36.7 days in the SSS-treated arm (n = 372), a 12.6-day difference (p = 5.5 × 10□□).

**Figure 4.**
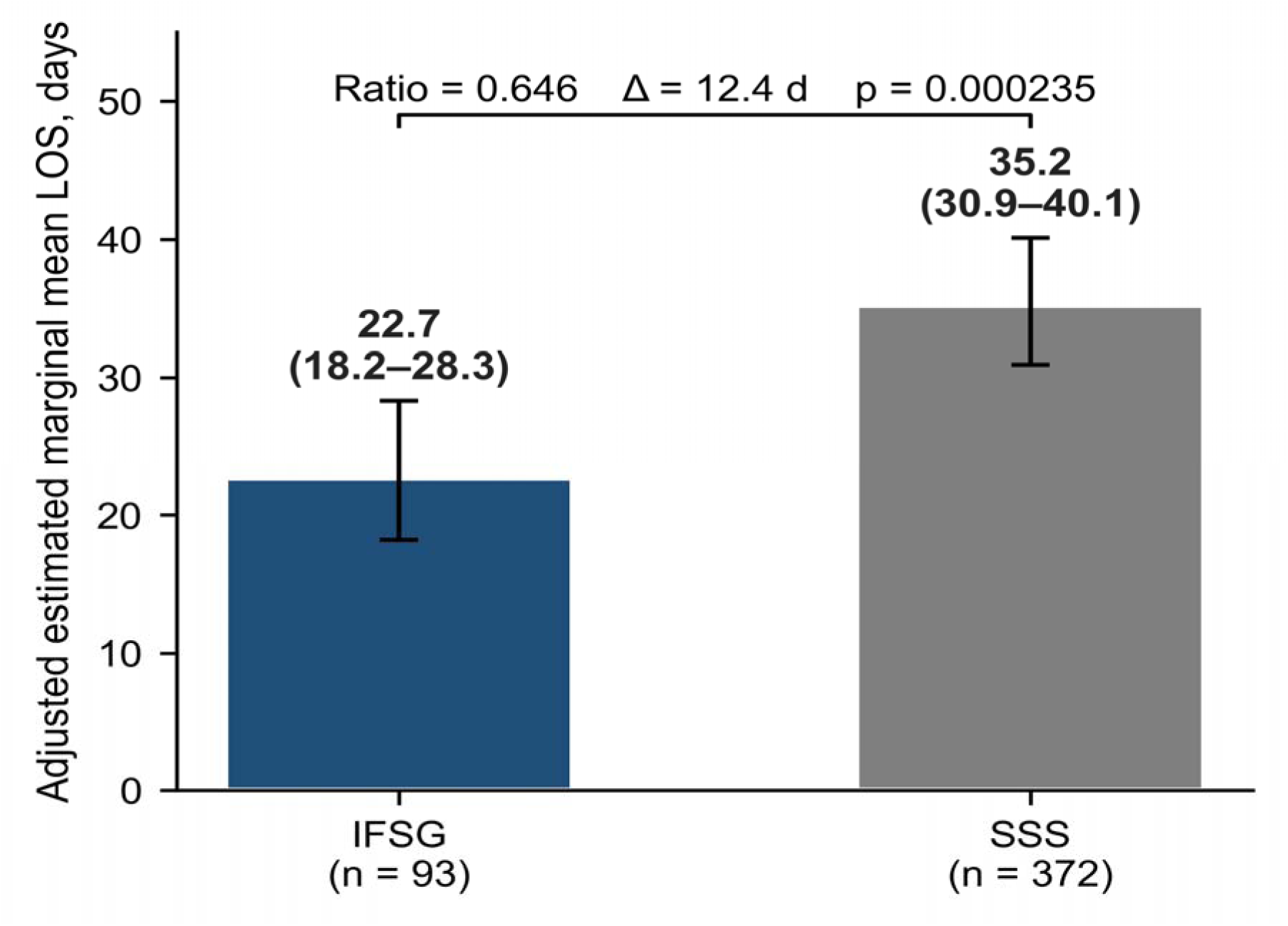
GLMM-adjusted estimated marginal mean (EMM) hospital length of stay with 95% confidence intervals (Gamma GLMM with facility and matched-set random effects). Adjusted LOS was 22.7 days (95% CI, 18.2–28.3) for IFSG versus 35.2 days (95% CI, 30.9–40.1) for SSS (ratio 0.646; p = 0.000235), a 12.4-day reduction in LOS for IFSG.

**Table 2.** Clinical Outcomes.

| Outcome | IFSG<br>(n = 93) | SSS<br>(n = 372) | Statistical test |
| --- | --- | --- | --- |
| **Primary Outcome** |  |  |  |
| LOS, mean ± SD (days) | 24.1 ± 17.4 | 36.7 ± 25.1 | p = 5.48e-08† |
| LOS, median [IQR] (days) | 20 [11–32] | 31 [17–52] | — |
| LOS, GLMM EMM [95% CI] (days) | 22.7 [18.2–28.3] | 35.2 [30.9–40.1] | p = 0.000235†† |
| **Secondary Outcomes** |  |  |  |
| Sepsis, n (%) | 1 (1.1) | 17 (4.6) | — |
| Graft loss, n (%) | 3 (3.2) | 31 (8.3) | — |
| VTE, n (%) | 2 (2.2) | 10 (2.7) | — |
| HAPI, n (%) | 2 (2.2) | 14 (3.8) | — |
| Any complication, n (%) | 7 (7.53) | 59 (15.86) | OR 2.31 (1.01–6.22), p = 0.045* |
| Composite, GLMM predicted % [OR 95% CI] | (6.73) | (14.60) | OR 2.37 (1.03–5.42), p = 0.041** |
| Composite, conditional logistic stratified on matched set | — | — | OR 2.37 (1.03–5.41), p = 0.042 |

Counts/percentages are from matched cohort data. Primary outcome LOS: † p-value, †† p-value
p-value from Gamma GLMM with facility + matched-set random effects (†). Composite secondary outcome p-values: \*
= Fisher's exact test (unadjusted); \*\* = binomial GLMM with matched-set random effects. 95% CIs in parentheses.
EMM = estimated marginal mean. IFSG = Kerecis Omega3 Burn. SSS = Integra DRT + NovoSorb BTM.

### Sensitivity Analysis Supports the Magnitude and Direction of the Difference in LOS

A mixed-cohort sensitivity analysis which included patients who received the index product alongside other dermal substitutes (n = 229 IFSG vs n = 458 SSS across 67 centers) yielded a GLMM-adjusted LOS of 27.6 days (95% CI 23.8–31.9) for IFSG versus 38.5 days (95% CI 34.8–42.6) for SSS (ratio 0.716; p = 0.0001; difference 10.9 days). The direction and magnitude of the LOS advantage in this broader patient population are concordant with the pure-cohort finding.

### Complication Rate Was Lower in the IFSG-Treated Group

Complication rates are presented in Table 2 and Figure 5. All four individual complications trended less common in the IFSG-treated group: sepsis (1/93, 1.1% vs 17/372, 4.6%), graft loss (3/93, 3.2% vs 31/372, 8.3%), VTE (2/93, 2.2% vs 10/372, 2.7%), and HAPI (2/93, 2.2% vs 14/372, 3.8%), individual comparisons were not statistically significant according to Fisher’s Exact Test (Figure 5). On the composite endpoint, 7/93 IFSG-treated patients (7.53%) experienced at least one complication compared with 59/372 SSS-treated patients (15.86%), an absolute difference of 8.33% that reached statistical significance by Fisher’s exact test (OR 2.31, 95% CI 1.01–6.22; p = 0.045). The primary GLMM with matched-set random effects estimated predicted complication probabilities of 6.73% in the IFSG arm versus 14.60% in the SSS arm, with an odds ratio of 2.37 (95% CI 1.03–5.42) for SSS versus IFSG (estimate = 0.862, z = 2.039, p = 0.041). A conditional logistic regression stratified on matched set, used as an additional robustness check, produced a concordant estimate (OR 2.37, 95% CI 1.03–5.41; p = 0.042). The convergence of three independent tests, unadjusted Fisher’s exact test, GLMM with matched-set random effects, and conditional logistic regression, supports the primary finding of a higher composite complication rate in the SSS arm.

**Figure 5.**
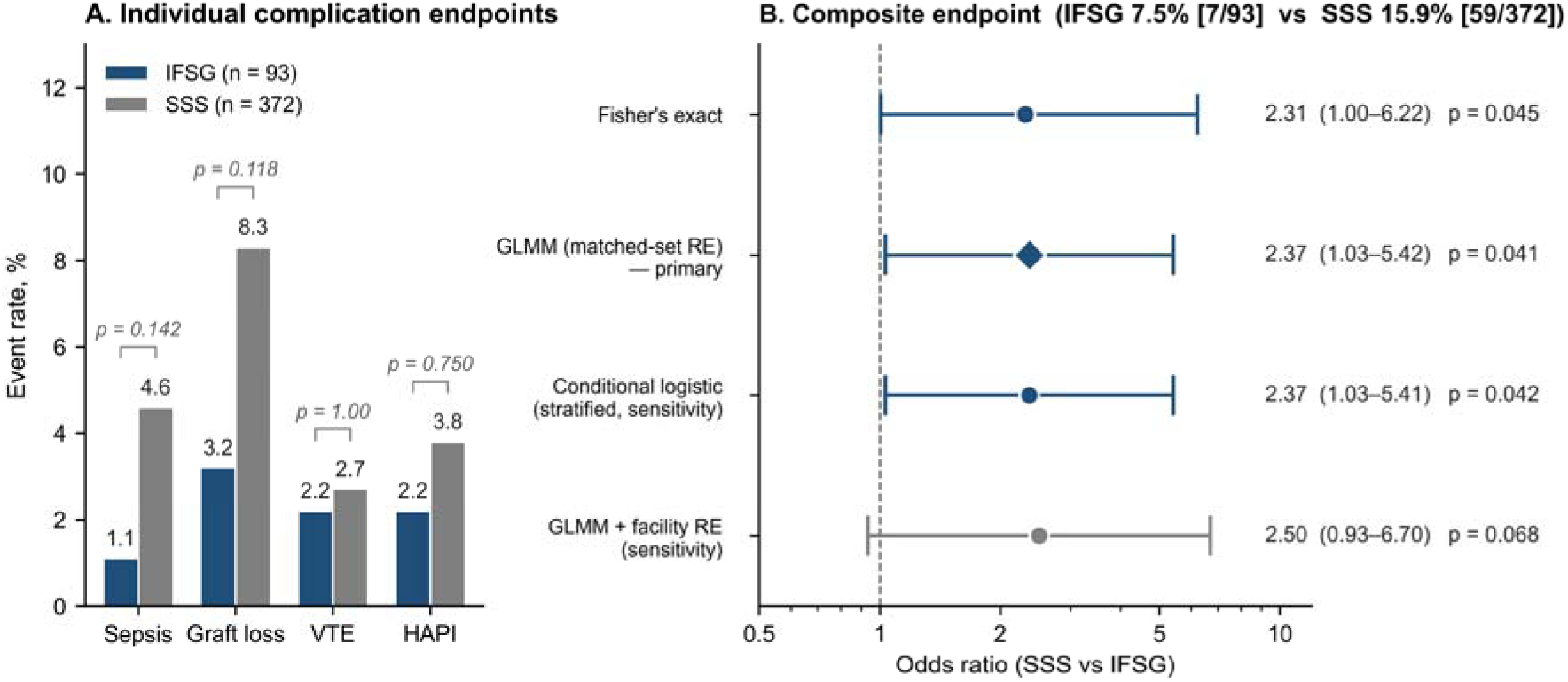
Complication outcomes for IFSG (n = 93) versus SSS (n = 372). (A) Raw event rates for the four individual complication endpoints (sepsis, graft loss, VTE, HAPI), each compared by Fisher’s exact test; all were numerically lower in the IFSG-treated arm but did not reach statistical significance. (B) Forest plot of odds ratios (SSS vs IFSG) for the prespecified composite endpoint (any of sepsis, graft loss, VTE, or HAPI). The primary GLMM with matched-set random effects gave OR 2.37 (95% CI, 1.03–5.42; p = 0.041); concordant estimates were obtained from Fisher’s exact (OR 2.31, 1.00–6.22; p = 0.045) and conditional logistic regression stratified on matched set (OR 2.37, 1.03–5.41; p = 0.042). The sensitivity model additionally including a facility random effect gave OR 2.50 (0.93–6.70; p = 0.068).

### Sensitivity Analysis Indicates the Difference in Complication Rate Should Be Confirmed With a Larger Sample Size

An additional sensitivity model added facility-level random effects to the primary matched-set structure. Under this model, predicted composite complication probabilities were 5.49% in the IFSG arm and 12.69% in the SSS arm (OR 2.50, 95% CI 0.93–6.70; estimate = 0.917, z = 1.824, p = 0.068). The effect size was preserved; the loss of statistical significance reflects the reduction in residual variance available for the fixed effect when partitioning variance across two clustering levels. Because the primary clustering structure for a 1:4 propensity-matched design is the matched set, the matched-set GLMM is the prespecified primary model; the additional facility-level model is presented as a robustness check. The lack of significance indicates the need for a larger sample size to confirm our results.

## Discussion

This study provides the first large-scale registry comparison of intact fish skin graft against established synthetic/biosynthetic dermal substitutes in a propensity score–matched burn population. After adjusting for facility-level clustering and matched-set correlation, IFSG was associated with a GLMM-adjusted estimated marginal mean LOS of 22.7 days compared with 35.2 days for SSS (ratio 0.646; p = 0.000235), a statistically significant and clinically meaningful difference of 12.4 days for burns of moderate total body surface area requiring dermal substitution and autografting. This reduction carries substantial implications for both patient outcomes and healthcare resource utilization.

Variables that have been shown to influence LOS include age, inhalation injury, TBSA, burn severity, trauma, and patient gender.^19–25^ A higher percent TBSA and older age are frequently identified as strongly predictive of a longer LOS.^19–21,23,24^ The presence of inhalation injury also predicts a longer LOS, as does the presence of concomitant trauma.^19,20,23^ Similarly, burns of greater severity (i.e. larger areas of full-thickness skin damage) are associated with a longer LOS.^22,24,25^ Patient gender has also been identified by studies as associated with LOS, though the direction of this association differs depending on the study.^22,23^ To reduce confounding by these variables known to predict LOS, the IFSG-treated and SSS-treated groups were matched for each of these covariates using propensity score matching. Diabetes was not included as a matching covariate because it is not among the established primary predictors of burn LOS.^20^ Notably, the IFSG arm carried a higher diabetes burden than the SSS arm (30.1% vs 21.2%), a recognized impediment to wound healing, yet IFSG-treated patients had shorter LOS and a lower composite complication rate, suggesting that the observed effect is robust to this unmatched confounder. Channeling bias, whereby surgeon preference might direct IFSG toward less severe burns, is also unlikely to explain the findings: prior to matching, the IFSG-eligible pool had a higher proportion of third-degree burns than the SSS pool (36.9% vs 26.0%). IFSG was therefore being used in equally severe burns as its synthetic counterparts.

### Complications

Complications remain a major contributor to prolonged LOS and adverse outcomes. Sepsis occurs in approximately 20% of adult burn patients and is associated with markedly increased mortality and complication rates.^21^ Burn wound infection and dysregulated inflammatory responses are key drivers of sepsis.^22^ In this study, sepsis rates were low in both cohorts, occurring in 1.1% of IFSG-treated patients and 4.6% of SSS-treated patients. Other complications, such as graft loss, which may occur in approximately 7–20% of cases depending on population and graft type, often necessitate reoperation and significantly extend LOS.^23–25^ Graft loss in this study was also relatively rare and occurred in 3.2% of IFSG-treated patients and 8.3% of SSS-treated patients. Venous thromboembolism (VTE), while less common (∼1.7% in published series), is associated with increased morbidity and longer hospitalization.^26^ VTE rates were at an acceptable level in both cohorts, at 2.2% and 2.7% in the IFSG-treated group and SSS-treated group, respectively. Hospital-acquired pressure injuries (HAPIs) are a serious and common complication of hospitalization.^27^ HAPIs are related to LOS bidirectionally; HAPIs increase LOS and longer LOSs are associated with a higher incidence of HAPIs.^28,29^ In this study, HAPI occurred in 2.2% of the IFSG-treated group versus 3.8% of the SSS-treated group.

The primary function of a dermal regeneration template is to prepare a viable wound bed that optimizes autograft take. A longer scaffold integration period does not inherently produce a better recipient site; it simply extends the time the patient potentially remains hospitalized and increases exposure to nosocomial infection and risk of thromboembolism and inpatient complications. The clinically relevant question is whether the wound bed produced by a given template supports successful graft take without excess complications. On this measure, IFSG performed favorably: every individual complication endpoint was numerically lower in the IFSG arm, and the composite endpoint reached statistical significance in both the unadjusted and primary GLMM analyses. IFSG achieved wound-bed preparation and definitive closure in a shorter timeframe while preserving the outcome that matters most to surgeons, reliable autograft take. The shorter hospitalization is therefore not merely a reflection of product design; it is itself the clinical advantage, realized alongside, rather than at the expense of, graft survival.

### Economic Implications

The economic implications of reduced LOS are substantial. LOS has been estimated to account for approximately 70% of total burn care costs,^35^ making it a critical driver of healthcare expenditure. In real-world analyses, even modest reductions in LOS have translated into significant cost savings; for example, a 3.3-day reduction in LOS with autologous skin cell suspension was associated with cost savings of approximately $36,949 per patient.^35^ Extrapolating from these data, the 12.4-day reduction observed in this study may correspond to substantially greater economic benefits, particularly in high-resource burn center settings.

### Limitations

This study has several strengths, including the use of a large national registry spanning 49 burn centers and robust propensity score matching to balance key confounders such as age, TBSA, and inhalation injury, variables consistently identified as major determinants of LOS. The analysis is structured around a single primary outcome and a single composite secondary outcome between two arms, which avoids the need for multiple-comparison correction and simplifies interpretation. However, limitations inherent to retrospective registry analyses must be acknowledged.

First, residual confounding may persist, and the registry lacked granularity regarding procedural timing and wound-specific characteristics. In particular, the matched cohort predominantly comprised medium-sized burns (median TBSA 13%); patients with extremely large burns requiring cultured epidermal autograft or multiple donor-site reharvesting cycles were underrepresented. In such cases, prolonged wound-bed staging may be clinically intentional, and the LOS advantage observed here may be attenuated. The sample size was insufficient for a meaningful subgroup analysis of large burns (≥40% TBSA). Lacey et al. reported that intermediate skin substitutes may be unnecessary in small burns affecting less than 10% of total body surface area, where sufficient donor skin is available and split-thickness autograft alone achieves satisfactory closure without the added cost and operative complexity of a two-stage procedure.^30^ The present cohort, with an average TBSA of 12.5%, spans a range from very small wounds where STSG alone may suffice to larger injuries where a dermal regeneration template provides clear clinical value. These findings should therefore be interpreted in the context of the burns for which a dermal template is indicated, principally deep partial- and full-thickness injuries of sufficient extent that wound-bed preparation and staged closure become necessary. For smaller, less complex burns, the LOS advantage observed here may be less clinically relevant, as the overall hospitalization is likely to be short regardless of product selection. This reinforces the importance of appropriate patient selection and suggests that the primary clinical benefit of IFSG may be most pronounced in medium-to-large sized burns requiring a dermal regeneration template.

Second, the SSS arm pooled IDRT and BTM patients as a single comparator group. This pooling was necessary because the registry only provides product-category level access for comparative analyses rather than individual product-level access. That is further justified by the shared mechanism of these products: both are bilayer dermal regeneration templates with a common surgical workflow (two-stage application with delayed autografting after vascular integration) and similar clinical indications for deep burns requiring wound-bed preparation prior to definitive closure.

Third, the registry does not capture wound-level data with sufficient granularity to control for wound location, surface area, or number of wounds treated. Fourth, while the pre-matching severity profile of the IFSG-eligible pool argues against channeling bias toward less severe burns depths, residual channeling effects related to other factors such as TBSA cannot be fully excluded; prospective randomized trials are needed to eliminate this concern definitively. Additionally, exclusion of cases with implausible LOS values was necessary to mitigate data recording errors but may introduce minor selection bias.

Finally, as a Kerecis-funded study, the potential for sponsor bias must be considered. We have mitigated this through independent data analysis by BData and an independent biostatistician, and by reporting all findings transparently.

## Conclusion

In burns requiring dermal substitution and split-thickness autografting, predominantly of moderate total body surface area, IFSG was associated with a statistically significant 12.4-day reduction in hospital length of stay and a statistically significantly lower composite complication rate compared with synthetic and biosynthetic dermal substitutes. The shorter hospitalization was therefore accompanied by reduced complications. Whether these advantages extend to massive burns requiring staged reconstruction, serial donor-site reharvesting, or cultured epidermal autograft remains to be determined in larger studies that allow for subgroup analysis. Given the substantial clinical and economic burden of prolonged hospitalization in burn care, IFSG is an effective dermal regeneration graft for improving both patient outcomes and healthcare efficiency. Prospective and larger studies are warranted to validate these findings and further define optimal patient selection and treatment pathways.

## CRediT Author Statement

Rajiv Sood: Conceptualization, Investigation, Validation, Writing – review & editing. Nathanael D. Hevelone: Methodology, Data curation, Formal analysis, Writing – review & editing. Ólafur B. Davíðsson: Methodology, Formal analysis, Writing – review & editing. Ragnar P. Kristjánsson: Methodology, Formal analysis, Writing – review & editing. Bart D. Phillips: Data curation, Project administration, Writing – review & editing. John C. Lantis II: Investigation, Validation, Writing – review & editing. Gunnar Johannsson: Conceptualization, Project administration, Writing – original draft, Writing – review & editing.

## Declaration of Competing Interest

RS and JCL have served as consultants to Kerecis. JCL is the Chairman of the Kerecis Scientific Advisory Board and the Director of their Clinical trials council; for which Little Black Raincloud Inc receives fair market compensation, JCL is a Co-Director of Little Black Raincloud Inc. NDH and BDP are employees of BData Inc., which received funding from Kerecis to perform the data extraction and propensity-matching analyses. ÓBD and RPK are employees of Frameshift ApS, which received funding from Kerecis to perform the independent statistical analyses. BData and Frameshift performed all analyses independently. The sponsor had no access to patient-level data at any point during the study. GJ is an employee of Kerecis (Coloplast), the manufacturer of the intact fish skin graft (GraftGuide) evaluated in this study.

## Funding

This study was funded by Coloplast Ltd (Minneapolis, MN, USA).

## Data Availability Statement

The data underlying this article were provided by the American Burn Association Burn Care Quality Platform (BData) under a data use agreement and cannot be shared publicly. Researchers may apply for data access directly through the American Burn Association / BData (https://ameriburn.org/bdata).

## Ethics Statement

Ethical review was performed by WCG IRB (reference 20261461), which determined the study exempt under 45 CFR 46.104(d)(4) for secondary analysis of de-identified data.

## Acknowledgments

The authors thank Sean Larson and Dan Schwartz (BData Inc.) for data management and operational support, and Mikkel Werling (Frameshift) for analytical assistance. The American Burn Association Burn Care Quality Platform provided the de-identified registry data used in this study.

## Notes

### Competing Interest Statement

This study was funded by Kerecis, LLC. RS and JCL have served as consultants to Kerecis. JCL is the Chairman of the Kerecis Scientific Advisory Board and the Director of their Clinical trials council; for which Little Black Raincloud Inc receives fair market compensation, JCL is a Co-Director of Little Black Raincloud Inc. NDH and BDP are employees of BData Inc., which received funding from Kerecis to perform the data extraction and propensity-matching analyses. OBD and RPK are employees of Frameshift ApS, which received funding from Kerecis to perform the independent statistical analyses. BData and Frameshift performed all analyses independently. The sponsor had no access to patient-level data at any point during the study. GJ is an employee of Kerecis (Coloplast), the manufacturer of the intact fish skin graft (GraftGuide) evaluated in this study.

### Funding Statement

This study was funded by Kerecis, LLC.

### Author Declarations

WCG IRB Connexus gave ethical approval for this work.

### Summary of Updates

The statistical analysis is now a two-arm analysis rather than a 3-arm analysis, better fitting the 2-arm comparison made in the paper. New analysis comparing number of patients experiencing at least one adverse event between the 2 arms has been added.

